# Classifying and visualizing medication use in the Adolescent Brain Cognitive Development (ABCD) Study

**DOI:** 10.1101/2025.11.19.25340321

**Authors:** Daniel A. Lopez, L. Nate Overholtzer, Kyung E. Rhee, Natalie Buchbinder, Gloria E. Ruiz-Orozco, Sam Steinhilber, Mia Tognoli, Arturo Lopez-Flores, Bonnie J. Nagel

## Abstract

**Background:** Medication use during adolescence provides important insight into current health and treatment patterns. However, these data are often difficult to analyze due to the complexity of medication labeling and classification. We present a reproducible framework that standardizes medication categorization in the Adolescent Brain Cognitive Development℠ Study (ABCD Study®), improving analytic consistency and enabling more reliable findings across the research community.

**Methods:** Parent-reported data on youth prescription and over-the-counter medication use from baseline through Year 6 of the ABCD Study were reviewed and harmonized. Medications were categorized using a combination of the Anatomical Therapeutic Chemical (ATC) Classification System, artificial intelligence–assisted methods, and expert clinical input. A case study illustrated the utility of this tool by examining longitudinal associations between antidepressant use and youth-reported internalizing symptoms on the Brief Problem Monitor (BPM-Y).

**Findings:** More than 6,300 unique medication entries were consolidated into 95 clinically meaningful categories, each coded across three recall periods (past 24 hours, past 2 weeks, past year). The most common unique medication labels were for cold/flu/allergy (553) and acne treatments (355). In the case study, adolescents taking antidepressants showed a significantly greater reduction in BPM-Y internalizing scores over time compared to nonusers.

**Conclusions:** This study introduces a standardized and reproducible classification of prescription and over-the-counter medication use in the ABCD Study. The resulting framework—accompanied by an interactive dashboard and publicly available code—facilitates new opportunities for researchers to examine how medication exposure relates to adolescent brain, neurocognitive behavior, and developmental outcomes.

## Introduction

Adolescence is a critical period of coordinated biological and neurodevelopmental change, during which many acute and chronic health conditions emerge, persist, or remit. Trends in medication use during this period offer valuable public health insight into shifting disease patterns and treatment practices. National surveillance data from 2011–2014 indicate that more than one in five youth in the United States used a prescription medication within the past 30 days, with notable shifts in prescribing patterns over the preceding decade (e.g., a decline in antibiotic use) (1, 2). More recent increases in antidepressant prescribing, particularly among adolescent females since the COVID-19 pandemic, underscore the importance of ongoing surveillance of medication use in youth (3).

At the individual level, medication exposure may serve as a confounder, mediator, or proxy for underlying health status. Incorporating medication use into population neuroscience and epidemiologic analyses is essential for accurate interpretation of adolescent developmental trajectories.

The Adolescent Brain Cognitive Development℠ Study (ABCD Study®) provides an unprecedented opportunity to examine these patterns, following more than 11,000 children enrolled at ages 9–10 years across 21 U.S. sites (4). Despite over 1,500 peer-reviewed papers using ABCD data, medication use remains infrequently included as an exposure or covariate. The caregiver-reported Medication Inventory Survey (MIS), part of the study’s physical health set of measures, records both prescription and over-the-counter products used (5). However, the MIS contains over 300 data fields per visit and lists medications using a mix of brand, generic, and formulation names—creating a substantial analytic barrier for researchers without pharmacologic expertise. In addition, medications are reported across three usage time frames (past 24 hours, past 2 weeks, past year), further complicating harmonization and longitudinal comparisons.

Mental health represents one important domain in which medication data are especially relevant to the ABCD Study. Psychiatric disorders affect nearly one in four U.S. youth by adulthood (Sappenfield 2024), and adolescence is a key period for the onset and persistence of psychopathology (6). At baseline, 9.7% of participants used at least one psychiatric medication per caregiver reported data (7), yet relatively few ABCD Study publications have examined medication use directly—most focusing on ADHD (8), and even fewer on non-psychiatric exposures (9, 10). Expanding analytic access to all medication types, including non-psychotropic and over-the-counter products, is essential to understanding the full spectrum of adolescent health and development.

Here, we describe a reproducible framework for categorizing medication data in the ABCD Study, and have created an interactive dashboard to increase usability of this data. The dashboard consolidates more than 6,000 unique prescription and over-the-counter medication entries into clinically meaningful categories. We summarize the prevalence of these categories across the baseline through Year 5 visits and the partial Year 6 visit, illustrating their application through a case study examining antidepressant use and internalizing symptoms. This framework is designed to enhance transparency, reproducibility, and accessibility for researchers investigating the diverse roles of medication use in adolescent development.

The ABCD dataset grows and changes over time. The ABCD data used in this report came from https://doi.org/10.82525/jy7n-g441. DOIs can be found at https://www.nbdc-datahub.org/abcd-release-6-0.

## Methods

The ABCD Study is a large, ongoing longitudinal cohort study of over 11,000 youth who were 9–10 years old at the baseline visit (11). Participants were recruited across 21 research sites in the U.S., along with a parent, caregiver, or legal guardian. Each annual visit includes a wide range of assessments, such as brain imaging, cognitive testing, mental and physical health outcomes, and a collection of demographic and behavioral information (5, 12).

### Medication Data

Medication use was assessed at every visit through the Medication Inventory Survey (MIS), completed by a parent or guardian. The MIS captures both prescription and over-the-counter (OTC) medications used in the past 24 hours and past 2 weeks. Medication use over the past year was also assessed beginning in Year 3. Parents or guardians were asked to bring medication containers to visits for verification. If the containers were not available, study staff followed up after the visit to collect the necessary information. Each medication was coded using a standardized RxNorm Concept Unique Identifier (RXCUI). No medication washout period was required as part of the ABCD protocol.

The current study includes data from baseline through Year 5 visits and partial Year 6 data (∼50% of the cohort). Only parent- or guardian-reported medications were included, covering both prescription and OTC medications.

### Mapping to ATC Classifications

We used the Anatomical Therapeutic Chemical (ATC) classification system (13) as the foundation for grouping medications reported in the ABCD Study. RxNorm identifiers were mapped to ATC codes using the Observational Health Data Sciences and Informatics (OHDSI) standardized vocabularies maintained by the OHDSI consortium (14–16). Mappings were drawn from the December 2024 OHDSI vocabulary release (ATHENA, https://athena.ohdsi.org/).

Medications with a single, unambiguous ATC match were directly assigned and manually verified. For medications with multiple plausible ATC matches, contextual information (e.g., common use patterns within the ABCD age range), artificial intelligence (AI)–assisted inference, and expert clinical input were used to determine the most likely classification. For medications without a definitive ATC match, categories were still assigned using complementary AI-based inference and expert consensus to approximate their probable therapeutic use. Overall, approximately 77% of unique RxCUIs had a direct ATC match; however, with these supplemental procedures, medication-category coverage was effectively complete, with nearly all medications assigned to a clinically meaningful group.

### Medication Categories

To enhance interpretability for researchers, we consolidated ATC codes into 95 medication categories, each represented by binary indicators for whether a participant reported use within a given timeframe (24 hours, 2 weeks, or past year). Past-year use was only assessed beginning in Year 3. Category definitions were tailored to pediatric and adolescent populations based on clinical relevance and use patterns. Sparsely populated categories that could not be merged were excluded. Final definitions were determined by author consensus and may be refined based on user feedback.

### Interactive Dashboard

To facilitate exploration of medication-use patterns in the ABCD Study, we developed an interactive dashboard using Tableau (version 2024.3.4), available at https://public.tableau.com/views/ABCD_Medications_v1/MedicationDashboard.

The dashboard contains a searchable database of all medications reported across study visits, along with each medication’s assigned Estimated Use Category and ATC classification. The Estimated Use Category represents a likely reason for use based on the medication’s common indication in populations similar to the ABCD sample and should not be interpreted as a definitive indication. To enhance transparency, each entry includes a tooltip listing additional possible uses.

The dashboard was generated exclusively from aggregate, de-identified data produced in R scripts. No participant-level records or identifiable information were used in any part of the dashboard; all visualizations summarize counts or percentages derived from pooled data across participants.

Users can search by medication generic or brand names (e.g., “Prozac” or “Fluoxetine”) to identify which specific medications were reported at least once through Year 6.

Longitudinal patterns of reported use can only be viewed at the medication-category level, rather than for individual medication names. Interactive filters allow users to focus on specific medication categories or timeframes (past 24 hours, past 2 weeks, or past year) and to compare patterns of category-level medication use across study visits.

The dashboard also includes interactive figures depicting the prevalence of medication use across time. For instance, users can select a category such as Antidepressant – SSRI to view the proportion of participants reporting use at each study visit, along with the underlying list of included medications. Multiple timeframes can be displayed side-by-side for comparison. Hovering over a data point reveals both the number of participants who endorsed taking a medication mapping to that category, the total number of participants that completed that ABCD Study visit, and the percentage of participants endorsing the medication category. For example, hovering over the Year 4 – Past Year data point for SSRIs displays that 622 participants reported use out of 9,714 who completed the Year 4 visit.

In addition to prevalence plots, the dashboard features a transition probability matrix illustrating the likelihood of continued use of a medication category across time, defined as P(Medication Use at Future Visit | Medication Use at Initial Visit). Rows represent the initial visit, and columns represent the future visit. For example, the probability of reporting Antidepressant – SSRI use at Year 3 among participants who reported use at Year 2 within the past 2 weeks was 0.77. Transition plots are currently limited to the 2-week and 24-hour timeframes, as past-year data were not collected before Year 3. Matrix cells may appear blank when probabilities could not be reliably estimated due to small sample sizes.

To prioritize participant privacy and data security, the released dashboard is restricted to displaying overall prevalence across study visits and transition probabilities. Demographic subgroup analyses and site-level breakdowns are not included.

### Statistical Analysis

The mapping of RXCUIs to ATC classifications was performed using SQL. Data processing and preparation for the dashboard were completed in R using RStudio and the Tidyverse package. (17–19).

### Case Study – Antidepressant Use and Youth Internalizing Scores

To illustrate the potential utility of this tool for ABCD Study researchers, we conducted a case study examining the association between antidepressant use and internalizing symptoms using the youth version of the Brief Problem Monitor (BPM-Y). The BPM-Y internalizing score is a count variable based on six items rated on a three-point Likert scale (0 = Not True, 1 = Somewhat True, 2 = Very True), with total scores ranging from 0 to 12 (20).

Because the distribution of internalizing scores was highly right-skewed, we fit a negative binomial mixed-effects model using the *glmmTMB* package (21). Covariates included household income, household education, participant age, and sex. We created a binary indicator of any antidepressant use in the past two weeks, combining SSRI and non-SSRI categories. An interaction between study visit and antidepressant use was tested to examine whether associations varied over time. Random intercepts for family ID nested within site accounted for the clustered ABCD design (22). The incidence rate ratio (IRR) with 95% confidence interval is reported as a measure of effect, and an IRR < 1.0 indicates the corresponding decrease in BPM-Y internalizing scores. A two-sided α of 0.05 was used to define statistical significance.

Example code for replication of the medication category definitions is available at https://github.com/Daniel-Adan-Lopez/ABCD_Medications.

## Results

There were 6,381 unique RXCUI–label combinations condensed to 95 medication categories. The most prevalent unique medication labels were cold/flu/allergy (553), acne treatments (355), ADHD medications (352), systemic antihistamines (340), and non-opioid pain treatments (324). Counts for the number of unique RXCUI-label combinations in each of the 95 medication categories are provided in Supplementary Table 1.

Table 1 presents participant characteristics stratified by whether participants ever reported medication use (including both prescription and over-the-counter medications) across any ABCD Study visit. Participants who ever reported medication use tended to be slightly older on average (13.1 vs 12.1 among non-users) and more likely to come from households with higher parental education and income. For instance, 37.4% of medication users had a parent with a graduate or professional degree compared to 31.8% among non-users, and over half (51.9%) of users were from households earning more than $100,000 annually, compared with 38.8% of non-users.

**Table 1.** Participant characteristics stratified by *ever* reporting medication use (prescription or over-the-counter) across any ABCD Study visit.

|  | Never reported medication<br>use (n=30,145) | Ever reported<br>medication use<br>(n=37,800) |
| --- | --- | --- |
| **Age, years, mean (SD)** | 12.06 (1.92) | 13.11 (2.03) |
| **Educational attainment of parents** |  |  |
| Up to high school (no diploma) | 1961 (6.6%) | 1031 (2.8%) |
| High school diploma/GED | 3456 (11.6%) | 2536 (6.8%) |
| Some college | 8462 (28.5%) | 10814 (28.9%) |
| Bachelor's degree | 6390 (21.5%) | 9002 (24.1%) |
| Graduate school or professional degree | 9438 (31.8%) | 13988 (37.4%) |
| **Household income** |  |  |
| < 50k | 8174 (27.4%) | 6675 (17.8%) |
| 50k to 100k | 7183 (24.1%) | 8717 (23.3%) |
| > 100k | 11562 (38.8%) | 19424 (51.9%) |
| Don't know | 1470 (4.9%) | 1478 (3.9%) |
| Refuse to Answer | 1396 (4.7%) | 1128 (3.0%) |
| **Race/ethnicity** |  |  |
| Hispanic | 7415 (24.6%) | 6205 (16.4%) |
| White | 13767 (45.7%) | 22826 (60.4%) |
| Black | 5411 (18.0%) | 4239 (11.2%) |
| Asian | 768 (2.5%) | 535 (1.4%) |
| Other/Multiracial | 2779 (9.2%) | 3991 (10.6%) |
| **Sex of child** |  |  |
| Male | 15712 (52.1%) | 19864 (52.6%) |
| Female | 14433 (47.9%) | 17936 (47.4%) |
| **BPM Youth Internalizing Score, mean (SD)** | 1.89 (2.3) | 2.32 (2.6) |

**Note.** Percentages reflect the proportion within medication-use groups, aggregated across all ABCD Study visits.

### Prevalence Across Time

Prevalence of medication use varied considerably across categories and over time, as shown in the Tableau dashboard (https://public.tableau.com/views/ABCD_Medications_v1/MedicationDashboard). For instance, SSRI use increased steadily from baseline, reaching 10.4% past-year prevalence at Year 6 (Figure 3). Any ADHD medication use showed differences depending on the recall window: at Year 4, 10.8% reported past-year use, 8.9% past two-week use, and 7% past 24-hour use. These differences underscore the importance of aligning the chosen time frame with the specific research question.

**Figure 1.**
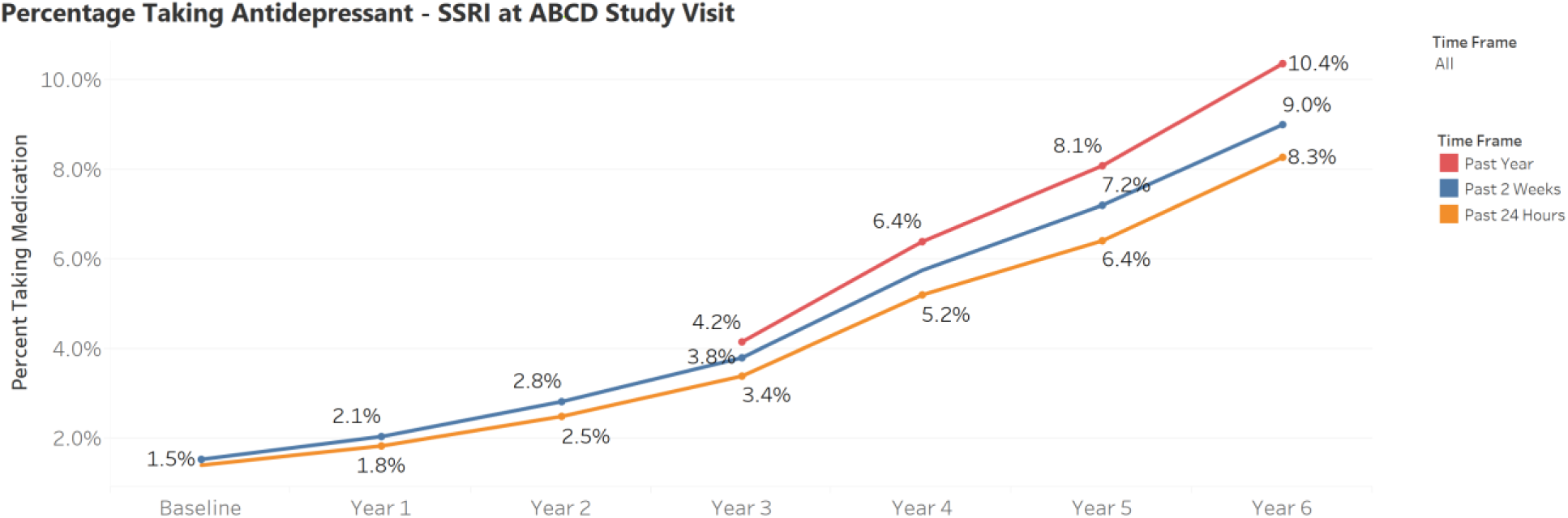
Percentage of participants reporting SSRI use at each ABCD study visit, by recall timeframe. Prevalence estimates are shown for use in the past year, past 2 weeks, and past 24 hours.

**Figure 2.**
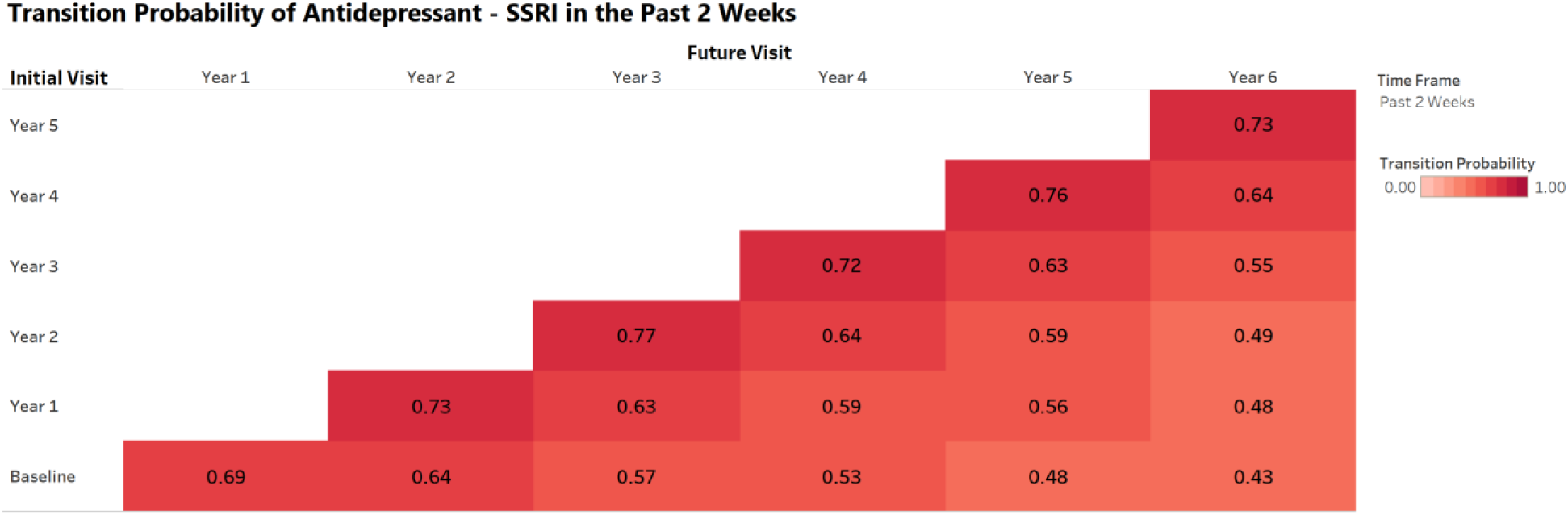
Example transition probability plot available in the medication dashboard. This visualization shows the probability of continued SSRI use across future ABCD study visits among participants reporting use in the past 2 weeks at a given initial visit. Darker shading reflects higher probabilities of sustained use.

**Figure 3.**
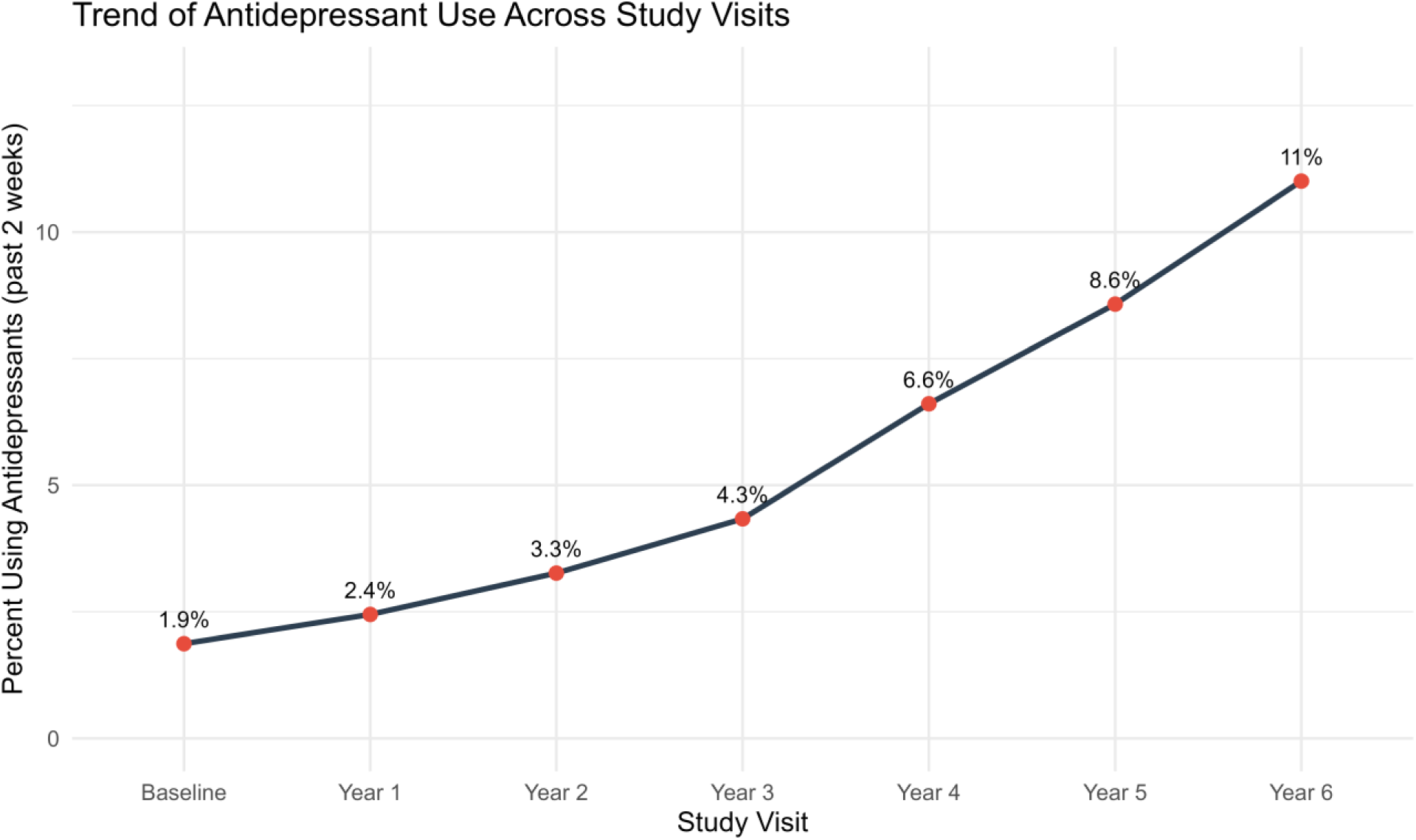
Prevalence of past 2-week any antidepressant use across ABCD study visits. The proportion of participants reporting any antidepressant use increased steadily from 1.9% at baseline to 11.0% by Year 6, indicating a growing prevalence of reported use over time.

**Figure 4.**
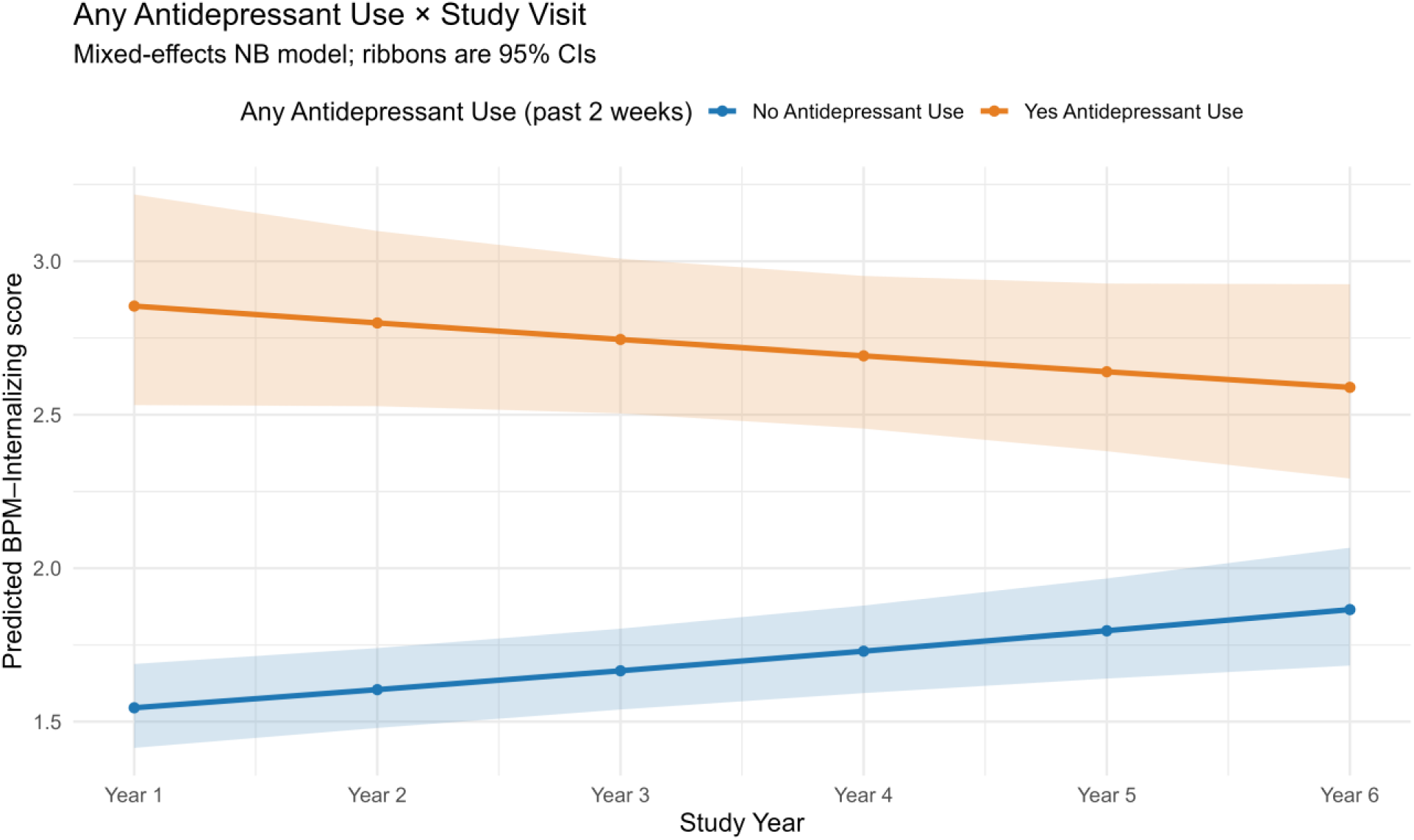
Interaction between any antidepressant use (past 2 weeks) and ABCD study visit on Brief Problem Monitor–Youth (BPM-Y) internalizing scores. Shaded ribbons represent 95% confidence intervals. An interaction effect indicated that internalizing symptoms increased less steeply across visits among youth reporting antidepressant use compared to non-users.

### Case Study – Use of Antidepressants and Child-Reported Internalizing Behaviors

The prevalence of past 2-week antidepressant use increased steadily across study visits, from 1.9% at baseline to 8.6% by Year 5, highlighting a growing proportion of participants reporting active antidepressant treatment over time (Figure 3).

In negative binomial mixed-effects models predicting internalizing symptoms (BPM–Y raw score), significant interactions between antidepressant use and study visit were observed for both the past 2-week and past 24-hour time frames (adjusted IRR = 0.94 [0.92, 0.97] and 0.95 [0.92, 0.97], respectively; *p* < .001) (Table 2). This indicates that, although antidepressant use became more common across study years, participants reporting use showed a slight decrease in internalizing scores compared to those not using antidepressants. In contrast, past-year antidepressant use was not significantly associated with change in internalizing symptoms over time (adjusted IRR = 0.98 [0.94, 1.02]; *p* = 0.38).

**Table 2.** Results of the interaction between ABCD Study Visit and Antidepressant Use on BPM-Y scores

| Predictor | BPM-Y Reported Score |  |
| --- | --- | --- |
|  | IRR (95% CI) | p value |
| Any antidepressant use past 2 weeks (ref = No) × study visit | **0.944** (0.92, 0.97) | p < .001 |
| Any antidepressant use past 24 hours (ref = No) × study visit | **0.946** (0.92, 0.97) | p < .001 |
| Any antidepressant use past year (ref = No) × study visit | 0.982 (0.94, 1.02) | p = 0.3772 |

**Note.** Negative binomial mixed-effects model with an interaction between antidepressant use (past 24 hours, past 2 weeks, or past year) and ABCD study visit. Results adjusted for age, sex, household income, household education, and a random intercept for family nested within study site. IRR = Incidence Rate Ratio; CI = Confidence Interval. An IRR < 1 indicates that internalizing scores increased less steeply among antidepressant users compared to non-users.

It is important to note that the “past year” medication-use variable was first administered beginning at Year 3, whereas the “past 24 hours” and “past 2 weeks” variables were collected starting at baseline. Consequently, estimates for the past-year exposure reflect a shorter follow-up window and are not directly comparable with the “past 24 hours” and “past 2 weeks” time frames.

## Discussion

This study introduces a reproducible framework for examining medication use in the ABCD Study by consolidating more than 6,000 unique RXCUI–label combinations into 95 clinically-relevant categories. This harmonization revealed meaningful differences in prevalence patterns both across medication classes and recall periods. Using a case study of antidepressant use and internalizing behaviors, we also demonstrated how medication categories can be leveraged to uncover clinically relevant associations in neurodevelopmental findings.

### Implications for Research

The development and open availability of reproducible medication categories and code greatly lower the barrier for studying medication use of any kind in the ABCD Study dataset. These resources will facilitate investigations into how psychotropic and non-psychotropic medications may influence adolescent development, including neuroimaging and neurocognitive outcomes, and temporal trends, including shifts during periods such as the COVID-19 pandemic.

Accounting for medication use is especially important in studies of brain–behavior relationships such as the ABCD Study, which did not include a washout period at any visit. Prior work illustrates how medication exposure can meaningfully alter observed associations. For example, a review of psychotropic medication effects in bipolar disorder found that antipsychotic use “normalized” structural MRI findings but not functional or diffusion measures (23), suggesting that medication exposure can mask true case–control differences. Similarly, a recent UK Biobank analysis found that analgesic and NSAID use were associated with better performance on multiple cognitive tests(24). Together, these findings emphasize that ignoring medication effects may introduce bias, particularly in neurocognitive and neuroimaging analyses.

### Case Study Findings

Our case study of antidepressant use illustrates how these categories can be applied to longitudinal data. Adolescents using antidepressants showed higher baseline BPM-Y Internalizing scores and a greater subsequent reduction in symptoms compared to those not taking antidepressants. These findings align with a meta-analysis of 15 randomized controlled trials (n > 12,000 adolescents) showing antidepressants were more effective than placebo in reducing depressive symptoms (25). While these findings are encouraging, they should be interpreted cautiously, as the ABCD Study represents an observational community-based sample.

### Confounding by Indication

Confounding by indication remains a critical consideration in all pharmacoepidemiologic research. In one national study, 27.5% of adolescents taking antidepressants met DSM-IV criteria for depression in the prior 12 months, compared with only 6.8% among non-users (26), reflecting approximately 5.7-fold higher odds of depression among those prescribed antidepressants. Such differences highlight why associations between medication use and symptom severity cannot be interpreted causally without accounting for baseline disease burden.

However, it is important to note that antidepressants are prescribed for a range of indications beyond major depressive disorder, including anxiety disorders, obsessive–compulsive disorder, and sleep or pain complaints. The term “antidepressant” therefore reflects pharmacologic class rather than clinical indication, complicating interpretation of exposure–outcome relationships. Confounding by indication occurs when the underlying reason for medication use (i.e., the clinical indication) is itself associated with the outcome (27). For example, adolescents with more severe internalizing symptoms are more likely to receive antidepressant treatment and more likely to continue exhibiting elevated symptom levels, potentially biasing naïve comparisons. While methods such as propensity score weighting, matching, or restrictive inclusion criteria can help address this bias (28), residual confounding is difficult to eliminate. Future ABCD Study analyses should explicitly address this issue when evaluating associations between medication use and youth outcomes.

### Strengths

This study has several strengths. First, it harmonized thousands of medication records for both prescription and over-the-counter medications into interpretable categories that can immediately support reproducible research within the ABCD Study. Medication data in large cohort studies are often underutilized due to the complexity of coding and labeling systems; our approach makes such data more accessible and easier to analyze. Although only 77% of medications received a direct ATC classification, we applied a combination of ATC mappings, AI–assisted inference, and expert input to assign categories for nearly 100% of all reported medications, providing a comprehensive foundation for future analyses. Second, the ABCD Study’s longitudinal design and demographic diversity make it uniquely suited for examining real-world medication use and its developmental correlates across adolescence. Third, by making all categorizations and accompanying code publicly available through an interactive dashboard and GitHub repository, we promote transparency, reproducibility, and ease of use. The inclusion of multiple recall periods (24 hours, 2 weeks, 1 year) further allows researchers to tailor their analyses to specific scientific questions and time frames of interest.

### Limitations

Several limitations warrant discussion. First, because indication data were not available, we cannot definitively determine why participants took each medication. Researchers using these categories should review them carefully and adapt classifications as appropriate. Second, the present work focused on parent-reported data, which may be subject to recall errors and underreporting. Child-reported medication data were first collected beginning at the Year 6 visit and will be incorporated in future updates as additional waves become available. Third, some medication categories have small sample sizes, which may limit statistical power for certain analyses. For privacy reasons, we excluded site- or demographic-level summaries from the public dashboard, and we encourage caution in interpreting results where counts are small. Additionally, medication use represents potentially sensitive health information, and researchers using this tool should adhere to ABCD Research Consortium privacy guidelines. Finally, the case study findings are illustrative rather than definitive. They demonstrate how to apply the categorization longitudinally, but are not designed as a primary research question. Future work should test alternative designs (e.g., within-subject or restricted cohorts) to more thoroughly investigate confounding by indication and its potential impact on observed associations.

### Future Directions

The ABCD Study provides an unprecedented opportunity to examine real-world patterns of medication use and their associations with adolescent brain and behavioral development. The standardized medication categories introduced here can be used to address a wide range of research questions. For instance, future work can investigate whether recent medication use (e.g., within 24 hours) influences neuroimaging or cognitive task performance, or how polypharmacy and cumulative exposure patterns relate to developmental trajectories of mental and physical health. Continued refinement of these medication categories, incorporating user feedback, will further enhance their accuracy and utility. Future updates will also include information on secondary indications and off-label use to provide a more comprehensive framework for pharmacoepidemiologic analyses within the ABCD Study cohort.

## Conclusion

This study provides a standardized, reproducible method for categorizing medication use in the ABCD Study and demonstrates its practical application through a case study of antidepressant use. These resources should enable researchers to explore a wide array of questions regarding medication exposure and adolescent development. Finally, this work underscores the importance of accounting for confounding by indication when interpreting associations between medication use and behavioral outcomes in observational data.

Data used in the preparation of this article were obtained from the <u>Adolescent Brain Cognitive Development℠ (ABCD) Study</u>, held in the NIH Brain Development Cohorts Data Sharing Platform. This is a multisite, longitudinal study designed to recruit more than 10,000 children age 9–10 and follow them over 10 years into early adulthood.

The ABCD Study® is supported by the National Institutes of Health and additional federal partners under award numbers U01DA041048, U01DA050989, U01DA051016, U01DA041022, U01DA051018, U01DA051037, U01DA050987, U01DA041174, U01DA041106, U01DA041117, U01DA041028, U01DA041134, U01DA050988, U01DA051039, U01DA041156, U01DA041025, U01DA041120, U01DA051038, U01DA041148, U01DA041093, U01DA041089, U24DA041123, U24DA041147. A full list of supporters is available at Federal Partners – ABCD Study.

A list of participating sites and a complete listing of the study investigators can be found on the Complete ABCD Study Roster. ABCD Consortium investigators designed and implemented the study and/or provided data but did not necessarily participate in the analysis or writing of this report. This manuscript reflects the views of the authors and may not reflect the opinions or views of the NIH or ABCD Consortium investigators.

## Data Availability

All data produced in the present study are available upon reasonable request to the authors.

---

**Table S1.**
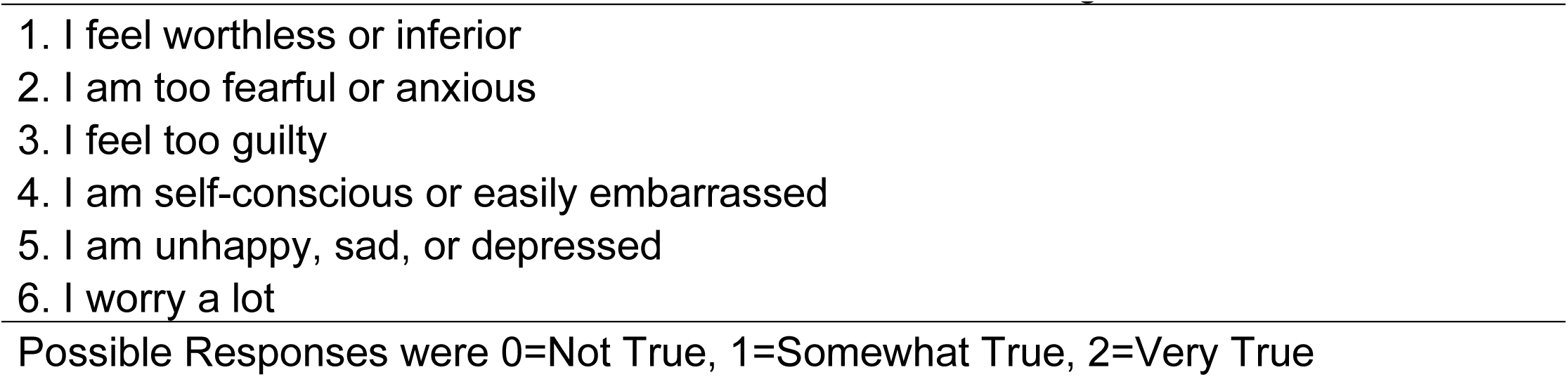
Items on the Brief Problem Monitor - Internalizing Scale

**Table S2.**
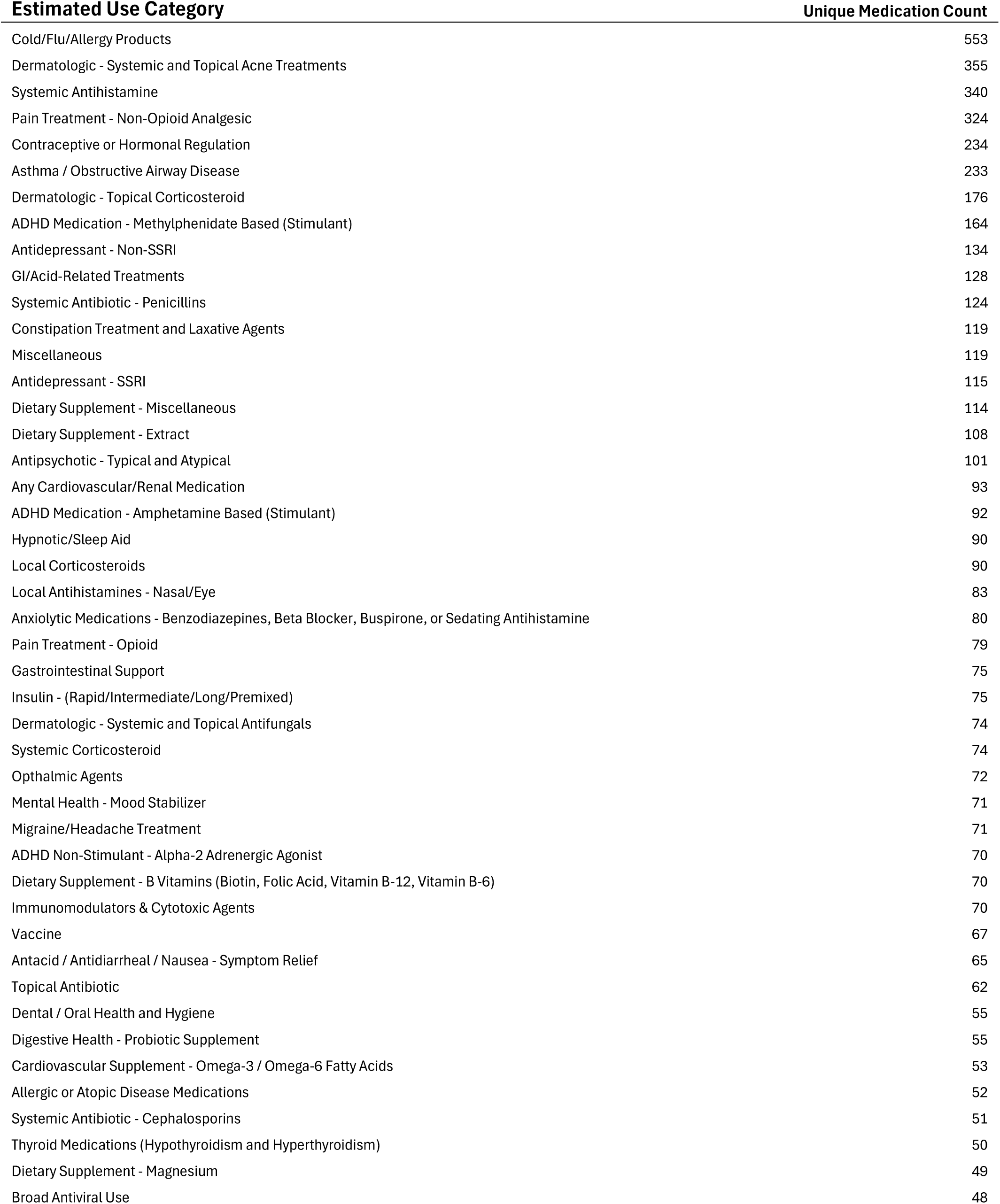
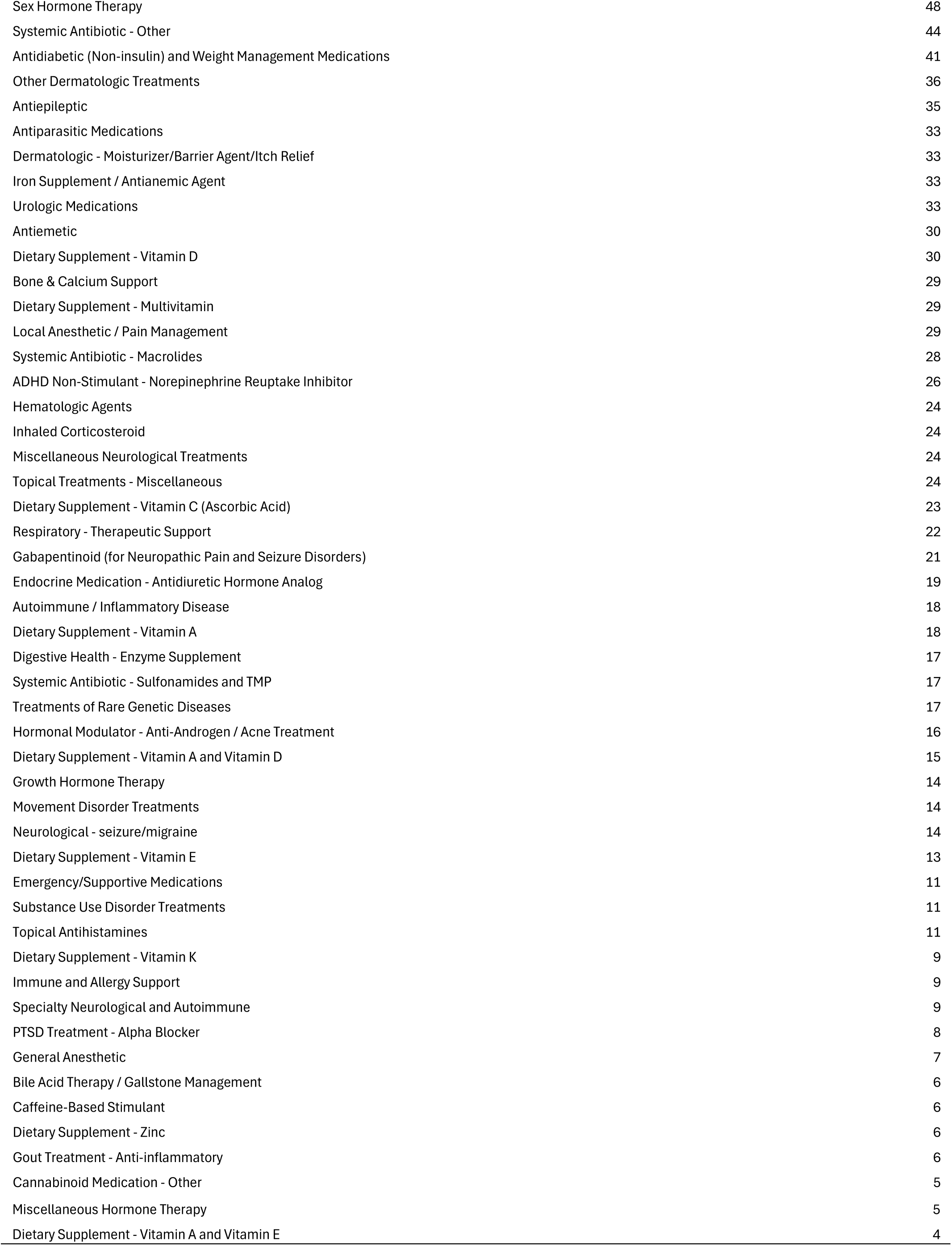

| **Estimated Use Category** | **Unique Medication Count** |
| --- | --- |
| Cold/Flu/Allergy Products | 553 |
| Dermatologic - Systemic and Topical Acne Treatments | 355 |
| Systemic Antihistamine | 340 |
| Pain Treatment - Non-Opioid Analgesic | 324 |
| Contraceptive or Hormonal Regulation | 234 |
| Asthma / Obstructive Airway Disease | 233 |
| Dermatologic - Topical Corticosteroid | 176 |
| ADHD Medication - Methylphenidate Based (Stimulant) | 164 |
| Antidepressant - Non-SSRI | 134 |
| GI/Acid-Related Treatments | 128 |
| Systemic Antibiotic - Penicillins | 124 |
| Constipation Treatment and Laxative Agents | 119 |
| Miscellaneous | 119 |
| Antidepressant - SSRI | 115 |
| Dietary Supplement - Miscellaneous | 114 |
| Dietary Supplement - Extract | 108 |
| Antipsychotic - Typical and Atypical | 101 |
| Any Cardiovascular/Renal Medication | 93 |
| ADHD Medication - Amphetamine Based (Stimulant) | 92 |
| Hypnotic/Sleep Aid | 90 |
| Local Corticosteroids | 90 |
| Local Antihistamines - Nasal/Eye | 83 |
| Anxiolytic Medications - Benzodiazepines, Beta Blocker, Buspirone, or Sedating Antihistamine | 80 |
| Pain Treatment - Opioid | 79 |
| Gastrointestinal Support | 75 |
| Insulin - (Rapid/Intermediate/Long/Premixed) | 75 |
| Dermatologic - Systemic and Topical Antifungals | 74 |
| Systemic Corticosteroid | 74 |
| Ophthalmic Agents | 72 |
| Mental Health - Mood Stabilizer | 71 |
| Migraine/Headache Treatment | 71 |
| ADHD Non-Stimulant - Alpha-2 Adrenergic Agonist | 70 |
| Dietary Supplement - B Vitamins (Biotin, Folic Acid, Vitamin B-12, Vitamin B-6) | 70 |
| Immunomodulators & Cytotoxic Agents | 70 |
| Vaccine | 67 |
| Antacid / Antidiarrheal / Nausea - Symptom Relief | 65 |
| Topical Antibiotic | 62 |
| Dental / Oral Health and Hygiene | 55 |
| Digestive Health - Probiotic Supplement | 55 |
| Cardiovascular Supplement - Omega-3 / Omega-6 Fatty Acids | 53 |
| Allergic or Atopic Disease Medications | 52 |
| Systemic Antibiotic - Cephalosporins | 51 |
| Thyroid Medications (Hypothyroidism and Hyperthyroidism) | 50 |
| Dietary Supplement - Magnesium | 49 |
| Broad Antiviral Use | 48 |
| Sex Hormone Therapy | 48 |
| Systemic Antibiotic - Other | 44 |
| Antidiabetic (Non-insulin) and Weight Management Medications | 41 |
| Other Dermatologic Treatments | 36 |
| Antiepileptic | 35 |
| Antiparasitic Medications | 33 |
| Dermatologic - Moisturizer/Barrier Agent/Itch Relief | 33 |
| Iron Supplement / Antianemic Agent | 33 |
| Urologic Medications | 33 |
| Antiemetic | 30 |
| Dietary Supplement - Vitamin D | 30 |
| Bone & Calcium Support | 29 |
| Dietary Supplement - Multivitamin | 29 |
| Local Anesthetic / Pain Management | 29 |
| Systemic Antibiotic - Macrolides | 28 |
| ADHD Non-Stimulant - Norepinephrine Reuptake Inhibitor | 26 |
| Hematologic Agents | 24 |
| Inhaled Corticosteroid | 24 |
| Miscellaneous Neurological Treatments | 24 |
| Topical Treatments - Miscellaneous | 24 |
| Dietary Supplement - Vitamin C (Ascorbic Acid) | 23 |
| Respiratory - Therapeutic Support | 22 |
| Gabapentinoid (for Neuropathic Pain and Seizure Disorders) | 21 |
| Endocrine Medication - Antidiuretic Hormone Analog | 19 |
| Autoimmune / Inflammatory Disease | 18 |
| Dietary Supplement - Vitamin A | 18 |
| Digestive Health - Enzyme Supplement | 17 |
| Systemic Antibiotic - Sulfonamides and TMP | 17 |
| Treatments of Rare Genetic Diseases | 17 |
| Hormonal Modulator - Anti-Androgen / Acne Treatment | 16 |
| Dietary Supplement - Vitamin A and Vitamin D | 15 |
| Growth Hormone Therapy | 14 |
| Movement Disorder Treatments | 14 |
| Neurological - seizure/migraine | 14 |
| Dietary Supplement - Vitamin E | 13 |
| Emergency/Supportive Medications | 11 |
| Substance Use Disorder Treatments | 11 |
| Topical Antihistamines | 11 |
| Dietary Supplement - Vitamin K | 9 |
| Immune and Allergy Support | 9 |
| Specialty Neurological and Autoimmune | 9 |
| PTSD Treatment - Alpha Blocker | 8 |
| General Anesthetic | 7 |
| Bile Acid Therapy / Gallstone Management | 6 |
| Caffeine-Based Stimulant | 6 |
| Dietary Supplement - Zinc | 6 |
| Gout Treatment - Anti-inflammatory | 6 |
| Cannabinoid Medication - Other | 5 |
| Miscellaneous Hormone Therapy | 5 |
| Dietary Supplement - Vitamin A and Vitamin E | 4 |

